# Gender attitudes, self-perceived traits, and developmental outcomes among Pakistani children in middle childhood

**DOI:** 10.64898/2026.01.27.26345000

**Authors:** Yulu Pan, Allison Frost, Lisa Bates, Aparna G. Kachoria, John A. Gallis, Victoria Baranov, Pietro Biroli, Joanna Maselko

## Abstract

This study examined gender attitudes and self-perceptions of culturally positive traits (bravery, leadership, and competitiveness) among 838 eight-year-old children (422 boys, 416 girls) in rural Pakistan. We assessed their attitudes patterns, and explored associations with mental health and academic outcomes. Overall, 35% of boys and 39% of girls attributed positive traits to both genders (egalitarian), associated with high self-perceptions of positive traits and more favorable outcomes. Children who endorsed gender stereotypes tended to favor their own gender, i.e. attribute positive traits only to their own gender. 5.5% of boys who attributed positive traits to women only (women-attributing) had lower Urdu (β= −0.50, 95% CI: −0.80, −0.20) and Math (β= −0.76, 95% CI: −1.06, −0.46) scores, while 12.7% of girls who attributed positive traits to men only (men-attributing) had modestly higher Math scores (β= 0.21, 95% CI: −0.03, 0.46). 9.5% of boys and 15.4% of girls attributed different positive traits to both genders (mixed-attributing), which was associated with poorer outcomes, including higher depressive symptoms among boys (β= 0.45, 95% CI: 0.09, 0.82). 36.0% of boys and 33.4% of girls reported high self-perception of positive traits, but self-perception alone was not strongly associated with outcomes. Findings suggest that, by middle childhood, children in rural Pakistan exhibit distinct gender attitudes that link with developmental outcomes. Notably, women-attributing and mixed-attributing attitudes were linked to poorer academic and mental health outcomes especially among boys, indicating potential educational and psychosocial costs of holding nonconforming gender views in patriarchal contexts.

## Introduction

Gender stereotypes, or generalized beliefs about individuals’ traits and behaviors based on their gender, can shape children’s self-perceptions, behaviors, and developmental outcomes (1,2). For example, girls who believe that boys are better at math may underestimate their ability and dedicate less time, effort, or attention towards math, resulting in poorer performance in those subjects. Research suggests that gender stereotype effects are context-dependent. This study provides evidence from Pakistan, a context where patriarchal norms and son preference remain dominant (3,4), but improvements in girls’ school enrollment and women’s labor force participation suggest gradual shifts in gender norms (5–7). In such patriarchal context with emerging egalitarian ideals, children’s gender attitudes (i.e. stereotypical or egalitarian attitudes about gender) are shaped by messages from multiple sources, and their attitudes may have implications for their mental health and academic performance (8,9). While most research in Pakistan has largely focused on adolescents and adults, little empirical research has examined gender attitudes during middle childhood (6 to 12 years old) in Pakistan, a critical period when children start to revise revise gender stereotypes with increasing flexibility and their own decisions can influence their behaviors and developmental outcomes (10,11). The current study addresses the gap by investigating gender attitudes and self-perception of positive traits among 838 eight-year-old children in rural Pakistan, as well as exploring their associations with developmental outcomes.

### Gender stereotypes across development

From early childhood, children are exposed to societal beliefs about traits and behaviors typically associated with a particular gender (2). They observe men and women occupying different roles, and receive either rewards for conformity or punishment for deviation from the dominant gender roles (12). These stereotypes often link masculinity to competitiveness, leadership, and dominance and femininity to warmth, nurturing, and empathy (8). Under patriarchal norms, men are expected to pursue paid work and social recognition which are more culturally valued, whereas women are expected to undertake caregiving and domestic tasks (8). Egalitarian norms, in contrast, challenge these rigid divisions and emphasize gender equality (13). By ages 3-4, most children are aware of concrete gendered expectations (e.g., dolls for girls and cars for boys), and these stereotypes tend to intensify through adolescence (2).

Across the developmental period from early childhood to adolescence, middle childhood (6 to 12 years old) is a transitional phase in which children can increasingly understand that men and women fulfill different roles (14). While gender stereotypes remain strong during middle childhood, children begin to understand counter-stereotypical examples, question the rigidity of traditional roles, and may revise their stereotypes flexibly (11,15,16). Cultural reinforcement during this period—through family, peers, media, and school—can either support or hinder the development of flexibility (14).

Evidence from adolescence shows that marked gender differences in stereotypical gender attitudes have already emerged (10). Across contexts, girls are generally less likely than boys to accept gender stereotypes, and boys’ gender attitudes are more strongly influenced by peer sanctions reinforcing masculine attributes (10). This pattern is observed in both high income countries and low- and middle-income countries (LMICs) (17,18). While most research of attitudes about gender targets adolescence, little is known about when these gender beliefs begin to diverge. Relative to the research on adolescence, empirical studies during middle childhood remain scarce (14,15), especially in South Asian settings where patriarchal norms and socialization processes may shape attitudes about gender from an early age (19).

### Gender attitudes and child outcomes

The extent to which children endorse gender stereotypes can shape their academic or vocational pursuits, as well as their socioemotional development and mental health(8,9). Numerous studies have demonstrated that children as young as six endorse pro-male stereotypes in STEM and pro-female stereotypes in verbal ability (11). Endorsement of such traditional gender stereotypes may restrict children’s self-efficacy (20) and lower their motivation and performance in areas that counter traditional expectations (21,22). Evidence from Europe suggests that egalitarian gender attitudes benefit all students’ performance in mathematics and reading (23,24). However, findings from China and Germany reveal context-specific and gender-specific effects. Some work shows that math-related gender stereotypes reinforce gaps in academic performance, with boys who endorse stereotypes performing better than girls (25). Other studies show that such stereotypes can motivate students to disprove expectations, leading to improved math performance among girls (26,28). These mixed findings suggest the academic impact of gender stereotypes may be moderated by the degree and type of stereotype endorsement, and broader socio-cultural context.

Across countries and age groups, egalitarian attitudes are generally associated with better mental health outcomes, while endorsement of traditional patriarchal beliefs (e.g., male dominance in decision-making) is associated with worse outcomes (29–33). However, holding egalitarian views can also be psychologically taxing when individuals who hold non-conforming attitudes encounter social pressure, discrimination and exclusion (8,34,35).These mixed findings underscore that while egalitarian attitudes are often beneficial for mental health, their impact depends on the surrounding social climate and the degree of support for gender equality.

### Gender attitudes and self-perception

Importantly, gender attitudes are often linked to children’s perceptions of their own gendered traits. Children may internalize gender stereotypes and perceive themselves in alignment with more masculine or feminine traits. Alternatively, children’s perceptions of their own attributes may shape how they view their gender more broadly, leading to their endorsement or rejection of gender stereotypes (36). Independent of attitudes about gender, children’s self-perceptions of gendered traits may impact their socioemotional outcomes. For instance, positive self-perceptions of masculinity traits, such as leadership abilities and competitiveness, are linked to lower depression risk (37), possibly through enhanced self-efficacy and active coping, which can buffer perceived stress and promote better mental health outcomes (38). In contrast, conformity to “toxic” masculine norms such as toughness and emotional stoicism can have harmful effects on mental health as they increase risk-taking behaviors and reduce willingness to seek social support (39). Examining children’s self-perceptions of gendered traits, especially traits that are culturally favorable, can inform our understanding of how children may internalize gender stereotypes and how gendered expectations may influence their development.

### The Pakistani context

In Pakistan, traditional norms often impose distinct expectations on men and women (3,4). In the patriarchal system, men typically serve as main breadwinners and decision-makers, while women are largely confined to domestic and caregiving roles with limited autonomy and decision-making power (3,40). Son preference remains prevalent, and parents strongly desire the birth of sons and prioritize educational investment in sons over daughters, with nearly half of Pakistani women lacking basic education (4,6).

However, recent decades have seen a gradual change. Girls’ school enrollment has steadily increased since the early 2000s (5,6), and women’s labor force participation has grown from around 14% in 2000 to 25% in 2020 (7). Gender equality movements and targeted interventions have expanded educational and economic opportunities for girls (41–44). While patriarchal attitudes persist, emerging survey evidence of Pakistani adults from middle-income households suggests increasing public acceptance of women’s participation in socio-economic domains, reflecting a gradual shift towards egalitarian norms (42).

### The Current Study

Measuring children’s gender attitudes and self-perception is important because these attitudes influence children’s beliefs and behaviors, ultimately shaping their future well-being and lifelong development. However, most research in Pakistan has largely focused on adolescents and adults, with limited evidence on younger children during middle childhood, when children start to develop gender flexibility and revise stereotypes (14,15). Existing studies also tend to focus narrowly on educational attainment and academic achievement (3,4,18,45), overlooking broader developmental outcomes.

This study addresses these gaps by investigating gender attitudes and self-perception of culturally positive traits—bravery, leadership, and competitiveness—among 838 children in middle childhood (8-years old) in rural Pakistan. Our goals are threefold: (1) to assess children’s gender attitudes and self-perceptions of positive traits, (2) to explore gender differences of their attitudes patterns, and (3) to examine how gender attitudes and self-perceptions are associated with developmental outcomes. This study contributes new evidence on the developmental implications of gender attitudes during middle childhood in a low-resource South Asian context.

## Materials and Methods

### Study population

This is a cross-sectional analysis of 838 children from the Bachpan cohort study, a birth cohort with a nested cluster randomized control trial evaluating a perinatal psychosocial depression intervention in Pakistan. The study design and characteristics of the Bachpan cohort have been described in detail elsewhere (47–49). Briefly, between October 2014 and February 2016, a total of 1,154 pregnant women in their third trimester living in 40 village clusters were recruited and screened for depression using the Patient Health Questionnaire-9 (PHQ-9), with a cutoff score of 10 or higher indicating depression (50). All these women were provided with a written consent form and consented volunarily for herself and her baby to participate in the study. All eligible women who screened positive for depression (N=570) were enrolled in a community-based trial of Thinking Healthy Program- Peer Delivered (THPP), a peer-delivered psychosocial intervention. One of every three women who screened negative for depression were enrolled in the cohort as non-depressed reference group (n = 584) (49). This study utilized data collection at baseline assessment (during pregnancy) and 8-year wave questionnaire. Of the 1,154 pregnant women enrolled at the baseline screening, 315 dyads were lost to follow-up at 8-year wave, and 1 additional dyad had missing data for the gender stereotype questionnaire and was excluded, leading to the final analytical sample of 838 dyads, including 422 boys and 416 girls (S1 Appendix). The Bachpan study received ethical approval from the institutional review boards (IRB) at the University of North Carolina at Chapel Hill, Duke University, the Human Development Research Foundation, and the National Bioethics Committee (Pakistan).

### Exposure Measures: Gender stereotyping and self-perception of positive traits

Informed by established gender-role attitudes research, the 8-year child questionnaire included items assessing both gender stereotypes and self-perception related to culturally valued and commonly studied traits in gender stereotype research: bravery, leadership, and competitiveness (1,51,52).

To assess gender stereotypes, Children were asked the extent to which they attribute the following three traits as typically male or female: *In your opinion, who can be brave/ a leader/ competitive?* Response options included “men only,” “women only,” or “men and women equally.” Based on their responses, children were categorized into one of four mutually exclusive Gender Stereotypes groups: 1) Egalitarian: answered “men and women equally” for all three traits; 2) Mixed-attributing: a combination of “men only” for at least one trait and “women only” for at least one trait; 3) Men-attributing: answered “men only” for at least one trait and never answered “women only”; 4) Women-attributing: answered “women only” for at least one trait and never answered “men only”. This classification allowed us to capture a child’s endorsement of traditional or egalitarian gender stereotypes, essential to understanding their self-perception in relation to these traits.

To assess self-perception, children were asked to what extent each trait described themselves: *How much do you think the following word describes you? Being brave / a leader/ competitive?* For each trait, children chose one of three response options: “0- Not at all”, “1-Somewhat”, “2- A lot”. Responses were summed to generate a total self-perception score, ranging from 0 to 6. The total scores were further classified into three levels to indicate the degree of self-perception: “high” (score = 6, 34.7%), “medium” (score = 4-5, 37.5%), and “low” (score = 0-3, 27.8%). The chosen cutpoints approximate the tertile grouping based on distribution of original score and allow for balanced group sizes.

### Developmental outcomes

Because gender attitudes and self-perception of positive traits may shape children’s emotional well-being, interpersonal interactions, and educational engagement, we examined multiple domains of child development to capture a broad range of potential impacts. We assessed five child outcomes at the 8-year wave involving domains of mental health, social-emotional development, and academic outcomes.

Mental health outcomes were measured by asking children to answer two established subscales of the Revised Child Anxiety and Depression Scale (RCADS)(53) and Spence Children’s Anxiety Scale (SCAS)(54). Specifically, we used major depressive disorder RCADS subscale (10 items, range 0-30) (55), and generalized anxiety SCAS subscale (6 items, range 0-18) (56), with higher scores indicating greater mental health burden. The Urdu versions of RCADS and SCAS have been used in Pakistani population with good internal consistency (Cronbach’s α > 0.85) (57,58).

Social and emotional outcomes were measured using the Strength and Difficulties Questionnaire (SDQ) (59), which has been previously validated for use in Pakistan (60). The SDQ questionnaire was completed by the mother based on her child’s behavior over the last six months or the current school year, and a Urdu version has been validated with good internal consistency (Cronbach’s α=0.7) in Pakistani population (61). The total score ranges from 0 to 40, and a higher score indicates more behavioral problems.

Child academic outcomes were assessed by asking children to complete the Urdu and Math test from Annual Status of Education Report (ASER), an assessment of children’s ability to read simple text and do basic arithmetic(62). The Urdu test score has 5 levels (0- Beginner level, 1- Alphabet level, 2- Word level, 3- Sentence level, 4- Story level). The Math test score has 6 levels (0- Beginner level, 1- Single digit level, 2- Double digit level, 3- Three digit level, 4- Subtraction level, 5- Division level).

### Confounders

We determined potential confounders a priori using a Directed Acyclic Graph (DAG) based on previous literature and knowledge(10,63). Most confounders were assessed at the 8-year wave, including mother’s mental health assessed with the PHQ-9 total score(50), index child’s trauma exposure in the previous year measured by the 15-item Traumatic Events Screening Inventory (TESI) (64), the level of environmental stimulation measured by the 15-item parental stimulation questionnaire (capturing the degree that any household member age 15 or over engaged in activities with index child in the past three days)(65), education aspiration (mother’s expectation of index child’s education years), mother’s and father’s education (total number of grades passed), child’s age in months, child’s grades (nursery, 1/2/3/4 grades, not in school), child’s school type (private school vs. other), and family structure (nuclear family vs. joint or multiple generation household). Maternal age during pregnancy, number of living children the mother has during pregnancy (first pregnancy, 1-3 children, 4 or more), and trial arm (control, intervention, non-depressed, based on the PHQ-9 cut-off and randomization at pregnancy), were measured at baseline. Household asset-based socioeconomic status (SES) quintile score (range 0-4, higher score indicates higher asset) (66), was assessed at the 6-year wave, which was the closest available data point.

### Statistical Analysis

Among the final analytical sample of 838 dyads, 15 had missing data for trauma exposure, 6 had missing data for education aspiration, 7 had missing for household asset-based SES quintile score. Given the low proportion of missingness, simple mean imputation was performed for these missing covariates. To account for informative missingness between baseline (N=1154) and 8-year wave (N=838), we used stabilized inverse probability of censoring weights (IPCW) so that non-missing individuals who share similar characteristics with missing individuals were upweighted in estimation. In the logistic regression model to calculate IPCW, we selected baseline variables associated with missingness (with the p-value threshold of 0.15) and baseline variables associated with child gender stereotypes and child outcomes (67). Selected baseline variables included maternal age, mother’s education, mother’s PHQ-9, trial arm, number of living children (first pregnancy, 1-3 children, 4 or more), family structure (nuclear family vs. joint or multiple household), household asset-based SES quintile score, life events and difficulties (LEDS) (68), maternal disability (WHODAS scale) (69), maternal perceived stress (70), crowding (number of people per room), total number of children, current major depressive episode, and husband’s working status (yes vs. no). The distribution of baseline variables used in IPCW modeling are presented in S5 Table.

For descriptive analysis of the core questions about the three positive traits (bravery, leadership, competitiveness), we calculated proportion of self-perception levels for positive traits (low/medium/high) within each gender stereotype group (egalitarian, mixed attributing, men attributing, women attributing) and compared the patterns among boys and girls.

All the child outcomes were Z-standardized for the primary analysis. To examine associations between gender stereotypes, self-perception, and child outcomes, we conducted separate linear regression models for 422 boys and 416 girls using generalized estimating equations (GEE), with children with “high” perception and children with “Egalitarian” gender attitudes as the reference group. Models were adjusted for all covariates described in the previous section. Robust standard errors accounted for the clustering assuming an exchangeable correlation structure, with individuals nested within villages.

To quantify the association between self-perception of traits, gender stereotypes, and child outcomes, the parameter coefficients (β) with 95% confidence interval (CI) for each group were estimated and interpreted as standardized mean differences. As a sensitivity analysis, complete case analysis was performed among 408 boys and 402 girls to compare with the primary results. All analyses were performed using SAS 9.4 (SAS Institute Inc., Cary, NC) and R 4.4.2 (R Foundation for Statistical Computing, Vienna, Austria).

## Results

Table 1 presents the characteristics of gender stereotypes, self-perception of positive traits, and child outcomes for 422 boys and 416 girls (Table 1). Overall, 35.1% of boys and 38.7% of girls held egalitarian gender attitudes, defined as attributing all three traits equally to men and women. Among children who expressed gender stereotypes, boys and girls tended to attribute positive traits more frequently to their own gender. Among boys, 50.0% attributed at least one of the traits to men and none to women (“men-attributing”), while only 5.5% attributed at least one of the traits to women and none to men (“women-attributing”). Similarly, but to a lesser degree, among girls, 33.2% endorsed women-attributing stereotypes, while only 12.7% endorsed men-attributing stereotypes. A subset of children – 9.5% of boys and 15.4% of girls – fell into the mixed-attributing group, defined as attributing traits to “men only” at least once and to “women only” at least once.

**Table 1.** Gender attitudes, self-perception, and outcomes among 838 children.

| Child Characteristics | Boys (N=422) | Girls (N=416) | p-value <sup>h</sup> |
| --- | --- | --- | --- |
|  | Mean ± SD / n (%) |  |  |
| Child age in months | 96.8 ±1.02 | 96.8 ±1.08 | 0.71 |
| School grades | 2.01 ±0.97 | 2.34 ±0.90 | <0.001 |
| Gender attitudes group <sup>a</sup> |  |  | <0.001 |
| Egalitarian | 148 (35.1%) | 161 (38.7%) |  |
| Mixed-attributing | 40 (9.5%) | 64 (15.4%) |  |
| Men-attributing | 211 (50.0%) | 53 (12.7%) |  |
| Women-attributing | 23 (5.5%) | 138 (33.2%) |  |
| Self-perception score | 4.4 ±1.6 | 4.4 ±1.6 | 0.69 |
| Self-perception group <sup>b</sup> |  |  | 0.44 |
| Low (0-3) | 114 (27.0%) | 119 (28.6%) |  |
| Medium (4-5) | 156 (37.0%) | 158 (38.0%) |  |
| High (6) | 152 (36.0%) | 139 (33.4%) |  |
| Self-perception of “a lot” for being brave | 248 (58.8%) | 230 (55.3%) | 0.31 |
| Self-perception of “a lot” for being a leader | 215 (50.9%) | 220 (52.9%) | 0.57 |
| Self-perception of “a lot” for being competitive | 307 (72.8%) | 292 (70.2%) | 0.41 |
| Developmental outcomes |  |  |  |
| Major depressive disorder (RCADS) <sup>c</sup> | 4.5 ±4.1 | 4.4 ±4.2 | 0.55 |
| Generalized anxiety (SCAS) <sup>d</sup> | 4.2 ±3.4 | 5.0 ±3.8 | 0.001 |
| Strengths and difficulties score (SDQ) <sup>e</sup> | 13.3 ±4.8 | 12.8 ±4.9 | 0.11 |
| Urdu test score <sup>f</sup> (mean) | 2.3 ±1.4 | 2.9 ±1.3 | <0.001 |
| Beginner level | 33 (7.8%) | 14 (3.4%) |  |
| Alphabet level | 105 (24.9%) | 68 (16.3%) |  |
| Word level | 112 (26.6%) | 81 (19.5%) |  |
| Sentence level | 36 (8.6%) | 36 (8.6%) |  |
| Story level | 135 (32.0%) | 217 (52.2%) |  |
| **Math test score <sup>g</sup> (mean)** | 3.3 ±1.3 | 3.5 ±1.2 | 0.02 |
| Beginner level | 14 (3.3%) | 5 (1.2%) |  |
| Single digit level | 38 (9.0%) | 27 (6.5%) |  |
| Double digit level | 78 (18.5%) | 82 (19.7%) |  |
| Three digit level | 30 (7.1%) | 16 (3.9%) |  |
| Subtraction level | 201 (47.7%) | 211 (50.7%) |  |
| Division level | 60 (14.3%) | 75 (18.0%) |  |

Descriptive statistics are presented as mean ± SD for continuous variables and n (%) for
categorical variables.
<sup>a</sup>. Gender at group were mutually exclusive based on how children attribute three traits to
male/female or both genders: 1) Egalitarian: answered “men and women equally” for all three
traits; 2) Mixed-attributing: a combination of “men only” for at least one trait and “women only”
for at least one trait; 3) Men-attributing: answered “men only” for at least one trait and never
answered “women only”; 4) Women-attributing: answered “women only” for at least one trait
and never answered “men only”.
<sup>b</sup>. The total self-perception score is a sum of responses to how much each trait described

Boys and girls showed similar patterns of self-perception across three positive traits. (Table 1) Over one-third of children (36.0% of boys and 33.4% of girls) reported high self-perception, defined as choosing “a lot” to describe themselves for all three traits (brave, leader, competitive). Meanwhile, 27.0% of boys and 28.6% of girls were categorized as having low self-perception of positive traits. Looking at each trait individually, slightly more than 50% of both boys and girls reported that the traits “brave” and “leader” described them “a lot”, and about 70% of both genders for the trait “competitive.” These results suggest that the majority of boys and girls in our sample are self-confident, and that gender does not play a major role in children’s self-perception.

Considering the relation between gender stereotypes and self-perception levels, Fig 1A and Fig 1B display the proportion of the three self-perception levels and mean score within each gender stereotype group, stratified by gender. Among boys, those with egalitarian gender attitudes had the highest level of self-perception, with the highest mean score of 4.6 and 43.9% classified as having high self-perception. Boys in the mixed-attributing group showed low self-perception of positive traits in general (mean score 3.5), with only 15% in the high self-perception category and 52.5% in the low self-perception category. Among girls, those with women-attributing attitudes had the highest self-perception of positive traits (mean score of 4.7, 42.8% having high self-perception), followed by those with egalitarian gender attitudes (mean score 4.5, 39.1% having high self-perception). Girls in mixed-attributing and men-attributing groups had lower self-perception, with approximately 15% in the high category and around 40% in the low category. Disaggregating these results for each trait, S1 – S3 Fig presents the percentage of self-perception levels for each trait within the trait-gender attribution response.

**Fig 1A.**
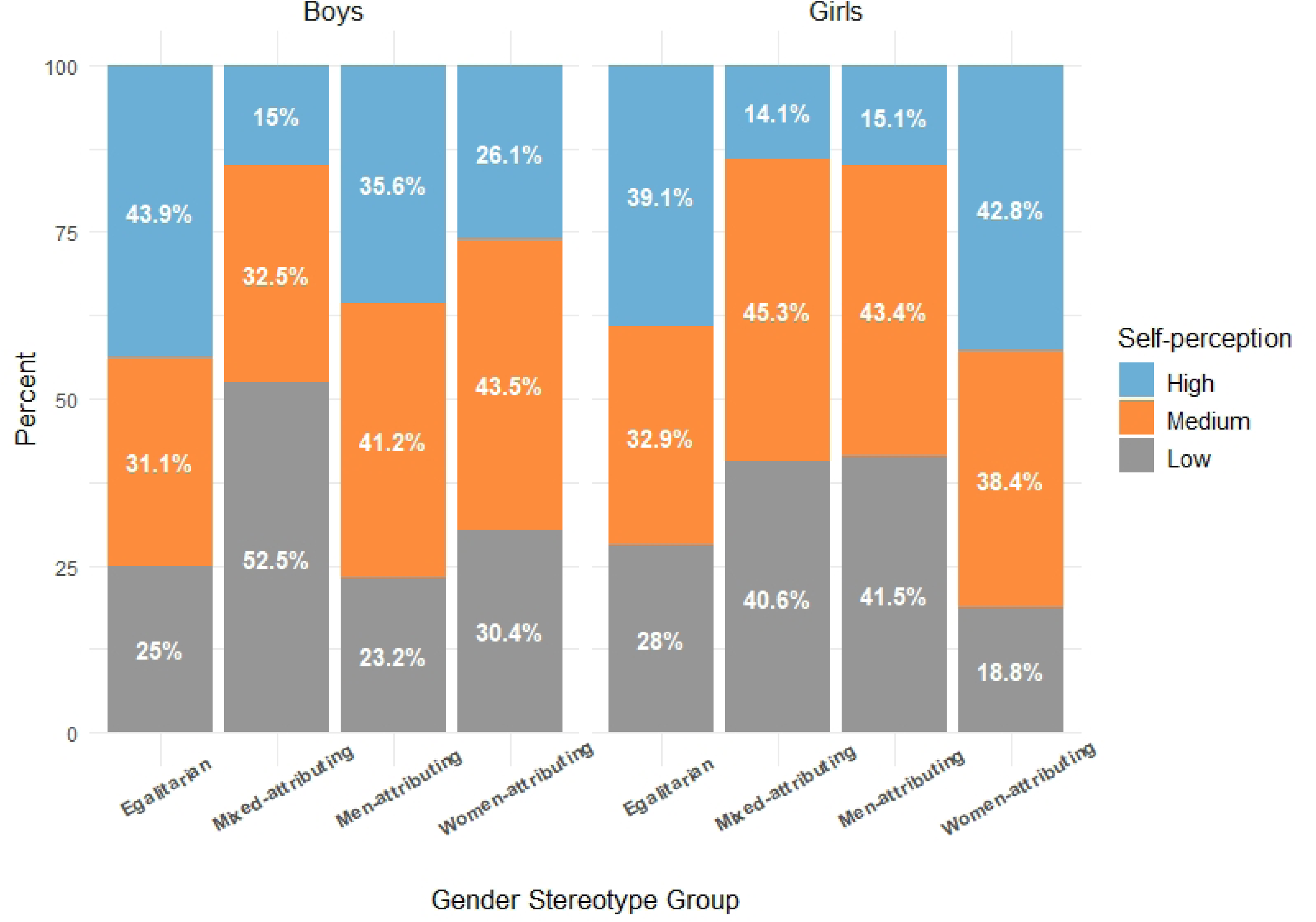
Proportion of self-perception levels for boys and girls within each gender stereotype group. Y axis indicates the proportion of self-perceptions levels (high/medium/low) within each of four gender gender stereotypes groups among boys and girls. Three self-perception levels according to the total self-perception score include: “high” (score = 6), “medium” (score = 4-5), and “low” (score = 0-3). Four mutually exclusive gender stereotypes groups include: 1) Egalitarian: answered “men and women equally” for all three traits; 2) Mixed-attributing: a combination of “men only” for at least one trait and “women only” for at least one trait; 3) Men-attributing: answered “men only” for at least one trait and never answered “women only”; 4) Women-attributing: answered “women only” for at least one trait and never answered “men only”.

**Fig 1B.**
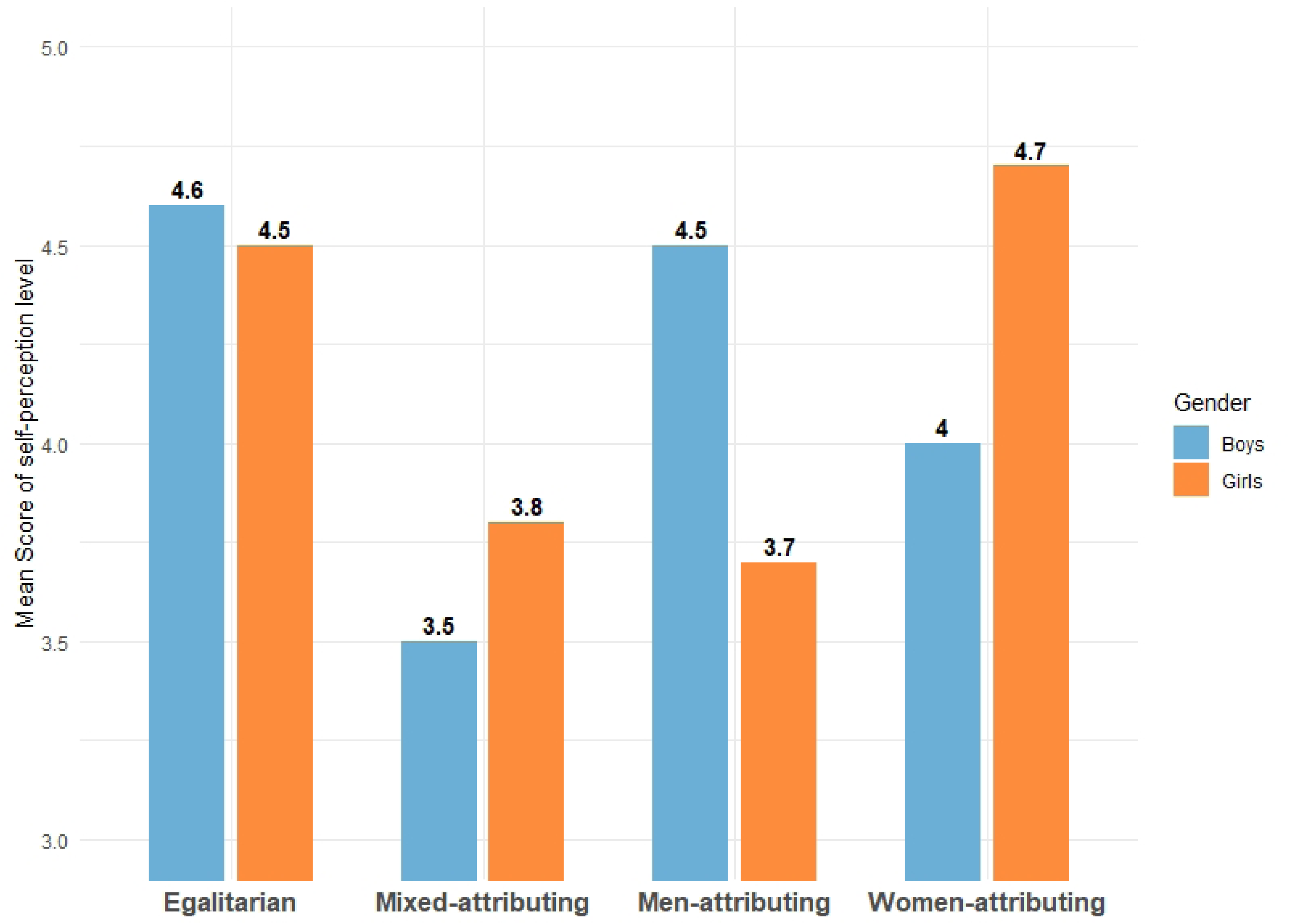
Mean self-perception score for boys and girls within each gender stereotype group. Y axis indicates the mean of total self-perception score within each of four gender gender stereotypes groups among boys and girls. Four mutually exclusive gender stereotypes groups include: 1) Egalitarian: answered “men and women equally” for all three traits; 2) Mixed-attributing: a combination of “men only” for at least one trait and “women only” for at least one trait; 3) Men-attributing: answered “men only” for at least one trait and never answered “women only”; 4) Women-attributing: answered “women only” for at least one trait and never answered “men only”.

In terms of developmental outcomes, girls demonstrated higher anxiety levels and higher Urdu and Math test scores than boys (Table 1). The distribution of other covariates during pregnancy (baseline) and 8-year wave are presented in S1 Table. Endorsed gender stereotypes showed varied associations with multiple child outcomes, and these associations differed by gender. Fig 2 and S2 Table present the adjusted coefficients of standardized child outcomes by gender attitudes groups, using the “Egalitarian” attitude as the reference. Mixed-attributing stereotypes were associated with negative mental health outcomes especially among boys, and the association between mixed-attributing attitude and depressive symptoms was significant (β= 0.45, 95% CI: 0.09, 0.82) among boys. The clear patterns emerged when comparing own-vs. other-gender attributions. Among boys, the associations between men-attributing and all the outcomes are close to the null value and not statistically significant. However, for boys, the direction of association between women-attributing and every outcome is deleterious and the inverse associations with the Urdu and Math outcomes are especially large and statistically significant (Urdu: β= −0.50, 95% CI: −0.80, −0.20; Math: β= −0.76, 95% CI: −1.06, −0.46). Among girls, the associations for stereotypical gender attitudes were mostly null, with the exceptions that girls with men-attributing attitudes had higher scores in Math (β= 0.21, 95% CI: −0.03, 0.46). Complete case analysis with 408 boys and 402 girls (S3 Table) produced similar results. Self-perception does not appear to be strongly associated with child outcomes for either boys or girls, as shown in Fig 3 and S4 Table.

**Fig 2.**
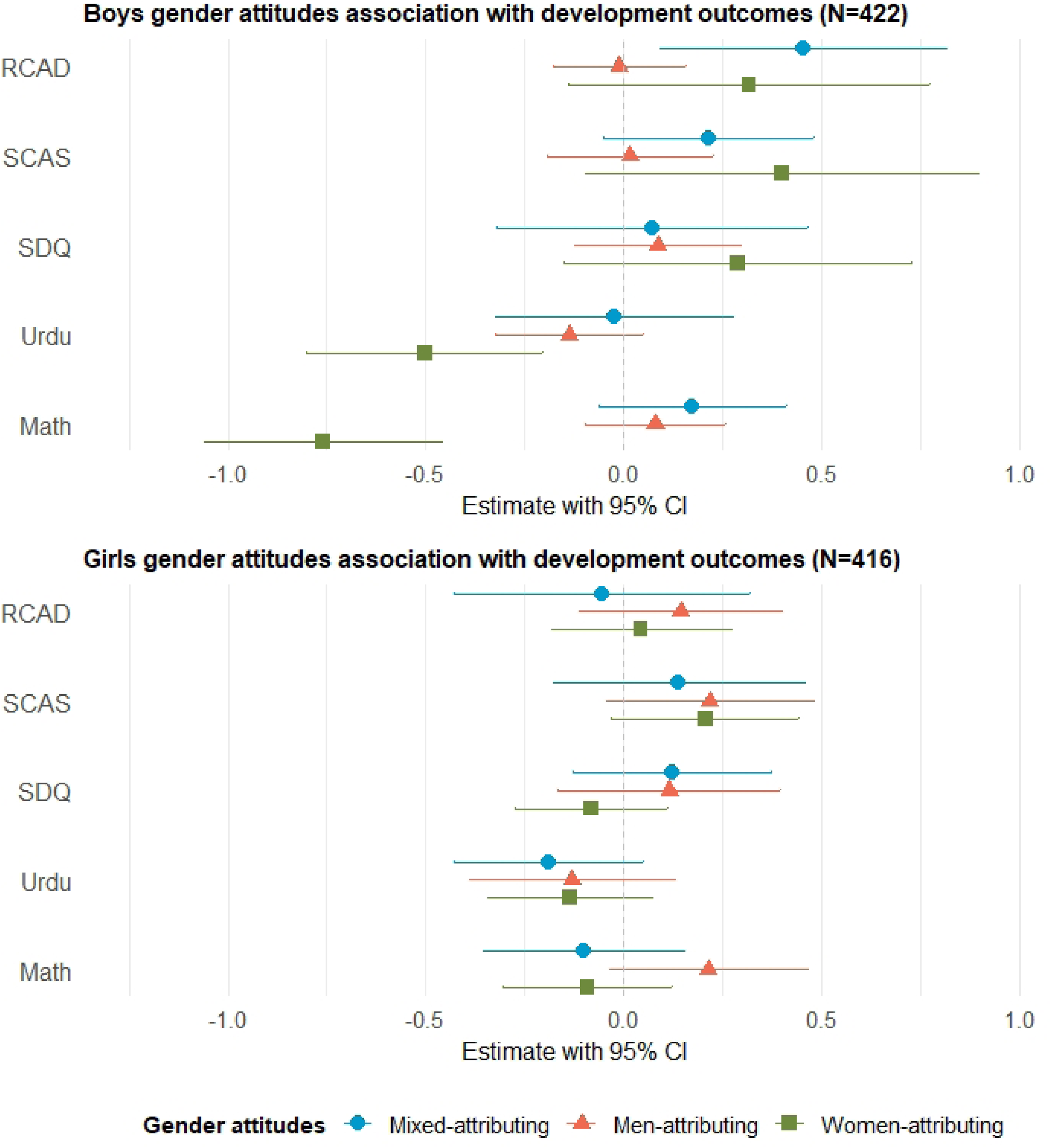
Boys and girls gender stereotype association with standardized outcomes. All the child outcomes were Z-standardized. The dots indicate the point estimate of association of between a particular gender attitude and child outcome, with egalitarian gender attitudes as reference group. The bars indicate 95% confidence interval. Abbreviations: RCAD = Major depressive disorder (RCADS); SCAS = Generalized anxiety (SCAS); SDQ = Strengths and difficulties score (SDQ); Urdu = Urdu test score; Math = Math test score. For outcome RACD, SCAS, SDQ, higher scores indicate worse outcome. For outcome Urdu, Math, higher scores indicate better outcome.

**Fig 3.**
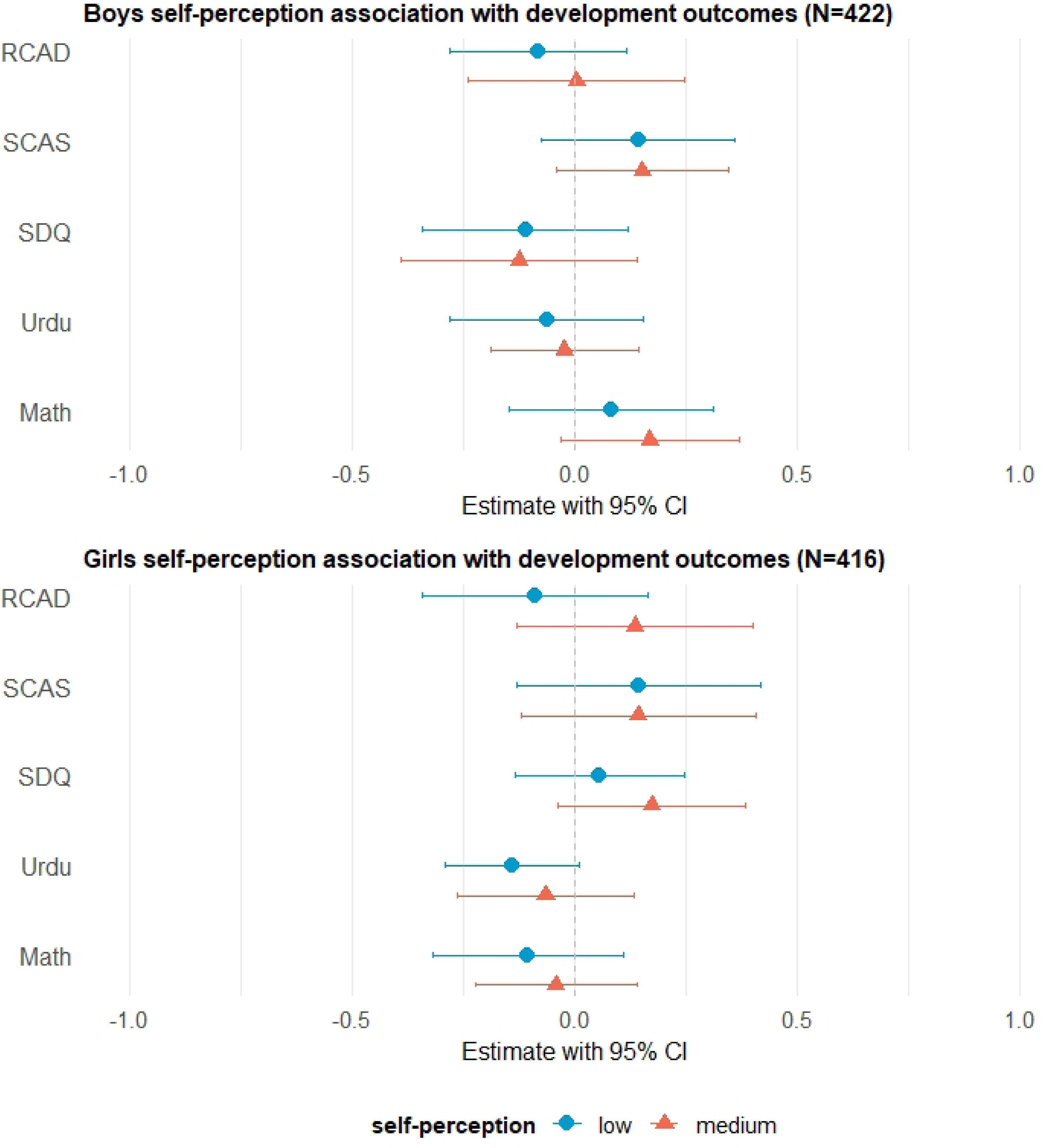
Boys and girls self-perception of traits association with standardized outcomes. All the child outcomes were Z-standardized. The dots indicate the point estimate of association of between self-perception level and child outcome, with high self-perception as reference group. The bars indicate 95% confidence interval. Abbreviations: RCAD = Major depressive disorder (RCADS); SCAS = Generalized anxiety (SCAS); SDQ = Strengths and difficulties score (SDQ); Urdu = Urdu test score; Math = Math test score. For outcome RACD, SCAS, SDQ, higher scores indicate worse outcome. For outcome Urdu, Math, higher scores indicate better outcome.

## Discussion

To our knowledge, this is the first investigation of how children express and potentially internalize gender attitudes, as well as perceive positive traits in their middle childhood in rural Pakistan. At age 8, children varied in the degree of self-perception of three culturally positive traits (i.e., bravery, leadership, and competitiveness), but the distribution did not differ notably by gender. By contrast, endorsement of gender attitudes of these traits varied considerably by gender. Girls were slightly more likely to hold egalitarian attitudes than boys, consistent with previous studies among adolescents and children in LMICs (17,18,71).

Erikson’s psychosocial development theory emphasizes that middle childhood is a crucial period for building a sense of self and competence, as well as being more aware of the evaluation from others (72,73). Differentiation of self-perception starts in early childhood, and continues to develop throughout adolescence and adulthood as individuals encounter new environments and social experiences (74). In our study, children in middle childhood showed differential levels of self-perception. Around 35% of children thought that they possessed “a lot” for all three positive traits, while 28% of children were considered as having low self-perception, indicating early variation in how children evaluate their own traits.

According to the gender schema theory, children internalize gender norms and apply them to both themselves and others (75). In patriarchal societies like Pakistan where men hold more power than women, positive traits such as bravery, leadership, and competitiveness are often culturally associated with masculinity (76). Thus, children exposed to such norms may begin to associate these positive traits more strongly with men than women. Indeed, we observed that boys were more likely than girls to attribute these traits to men (50.0% vs. 12.7%). Previous studies suggest that males are more likely to hold patriarchal beliefs that reinforce stereotypical gender roles (18,71,77). The findings also align with social dominance theory, which suggests that individuals from dominant social groups are more likely to support and internalize group-based hierarchies (78). In Pakistan, boys may benefit from adhering to dominant norms, including son preference, which affords them greater social value and access to resources (4). The dominant societal position of males may explain boys’ greater expression of patriarchal gender attitudes.

These patterns may not solely reflect conscious endorsement of gender stereotypes, especially among children in their middle childhood whose gender attitudes are still developing and revising. Instead, it may reflect “ingroup favoritism”, a common phenomenon that children have a strong bias favoring their own group (79). In our study, 33.2% of girls attributed these positive traits primarily to women, and 50.0% of boys attributed them primarily to men, supporting some degree of in-group favoritism. From a social identity theory perspective, attributing positive traits to one’s own group can reinforce positive group identity and self-esteem (80). Indeed, children who attributed positive traits to their own gender (i.e. boys with men-attributing and girls with women-attributing) had higher self-perception than those who attributed traits to the other gender. Such positive stereotype may be especially important for girls in patriarchal contexts, where girls need to shield themselves from internalizing social beliefs that undermine their capabilities and potential (81). Notably, girls with women-attributing attitudes had the highest level of self-perception, suggesting that holding positive stereotypes of female gender may serve as a protective mechanism for their self-esteem (9). Meanwhile, a small percentage of children attribute positive traits to the other gender and the pattern is more suggestive of internalized pro-male bias: the percentage of girls attributing the positive traits to boys was twice as high as that for boys attributing them to girls (12.7% vs. 5.5%).

Children in our study who held egalitarian gender attitudes also had relatively high self-perception of the positive traits. According to gender schema theory, children with more flexible gender attitudes are less likely to limit their self-perception to traits typically associated with their assigned gender, and more likely to internalize culturally valued traits regardless of gender (75). It is also possible that children with egalitarian attitudes have been exposed to more advanced education or have higher levels of cognitive development, both of which are linked to positive self-perception (82).

We also observed a unique mixed-attributing pattern, where children attributed different traits to both genders, though the mechanism is unclear and understudied. It may represent a transitional phase in gender attitude development, where children begin to challenge traditional gender stereotypes but have not yet developed a fully egalitarian attitude, or children receive traditional gender views towards men and women but have not yet developed a consistent stereotype. (15,16). According to social cognitive theory, children process and reconcile messages from various sources (83). When these messages are inconsistent, such as egalitarian attitudes at school but patriarchal beliefs at home, children may experience cognitive dissonance and find it hard to identify consistent role models and form coherent gender attitudes. Notably, children with mixed attributing attitudes had relatively lower self-perception of positive traits, possibly because the misalignment of cultural messages hinders internalization of culturally positive traits.

In terms of relationships between gender attitudes and child outcomes, while most adjusted associations were not statistically significant, one consistent patterns emerged: boys with women attributing attitudes, which is non-traditional in a patriarchal society, had significantly worse outcomes across multiple domains. Specifically, these boys exhibited higher levels of depression, anxiety, and behavioral difficulties, as well as lower academic performance in Urdu and Math tests compared to peers with egalitarian attitudes. Holding non-traditional attitudes in a patriarchal society can be mentally challenging, as individuals are more likely to experience peer pressure, social exclusion, and even emotional or physical violence (34), explaining worse mental health status for these boys. Favoring the other group also hinders confidence and self-esteem in their own group’s capabilities (80), which may reduce motivation and lower their academic performance.

Interestingly, girls with men-attributing attitudes had higher Math test score than those with egalitarian attitudes, which cannot be explained by in-group favoritism hypothesis. One possible interpretation is that some girls who attribute positive traits exclusively to men may still aspire to possess those traits, and set high achievement goals aligned with masculine ideals such as competitiveness. Alternatively, the pattern may also reflect internalized societal beliefs about male superiority in certain domains such as mathematics, which in turn motivates girls to work harder in order to parallel boys and disprove stereotypes. A similar pattern has been observed in China, where STEM-related gender stereotypes are widespread. Instead of being discouraged, many Chinese girls respond by increasing their effort and ultimately outperforming boys on math tests (26,28). This suggests that stereotype awareness does not necessarily diminish performance— and in some contexts may even drive compensatory motivation and higher achievement.

Unexpectedly, we did not find strong relationships between self-perception of positive traits and most child outcomes. This suggests that children’s beliefs about societal norms and group-based attributes may exert a stronger influence than internal self-views. In collectivist settings like rural Pakistan, where societal norms and expectations tend to be prioritized over personal beliefs (84), gender attitudes endorsement may play a more central role in shaping children’s well-being and achievement.

Our study has several strengths. First, it focuses on children in middle adulthood, a critical developmental stage for the formation of gender attitudes, in the rural Pakistan context that has been understudied. Second, we conducted a thorough investigation of the distribution of self-perception and gender stereotypes regarding three culturally positive traits, as well as their association with multiple child outcomes. Although the examination of these associations was exploratory, we applied rigorous statistical methods, accounting for clustering, informative censoring and potential confounding factors. Third, unlike most studies that dichotomize gender attitudes as either egalitarian or patriarchal, we captured the nuanced internalization patterns of gender stereotypes, including egalitarian, men-attributing, women-attributing, and mixed attributing. Fourth, all the analyses were stratified by gender, allowing us to compare gender differences and explore potential gender-specific mechanisms.

Nevertheless, we acknowledge some limitations. First, this is a cross-sectional analysis and longitudinal follow-up is necessary to assess how children’s gender attitudes evolve through adolescence and young adulthood. Second, the questionnaire only included three traits, which may not fully capture the breadth of individuals’ self-perception and gender trait attributions. Future studies can incorporate a wider range of traits, including traits stereotypically associated with femininity (such as nurturing and empathy), to provide a more comprehensive understanding of gender attitudes. Third, although the overall sample size was relatively large (n=838), some gender stereotype subgroups were small, leading to imprecise estimates. Lastly, we did not examine the reasons why children assigned certain positive traits exclusively to one gender, or assigned traits in a mixed way. Future qualitative or mixed-methods research would help unpack the underlying motivations and sociocultural contexts that shape gender attitudes. Understanding the reasons that contributed to their gender attitudes may illuminate the pathways through which gender stereotypes affect children’s development and well-being.

In conclusion, this study of 838 children in middle childhood in rural Pakistan revealed that a substantial proportion of both boys and girls reported high self-perceptions of positive traits, and endorsed egalitarian gender attitudes. Meanwhile, children at this age also begin to exhibit divergent patterns of gender attitudes. Importantly, certain gender stereotypes, particularly women-attributing and mixed-attributing patterns, were associated with poorer mental health and lower academic performance, with notable gender differences. Our findings suggest that in the context of rural Pakistan, children who held views contrasting with traditional patriarchal gender norms may experience internal conflict or social pressure. Future longitudinal and qualitative research is needed to explore how these attitudes evolve, and to inform interventions aimed at promoting gender equity and supporting healthy development of all children.

## Data Availability

Data will be made available on request.

## Author contributions

**Yulu Pan:** conceptualization, methodology, software, validation, formal analysis, investigation, writing - original draft, writing - review& editing, visualization.

**Allison Frost & Lisa Bates:** conceptualization, writing - review & editing.

**Aparna Kachoria &Victoria Baranov & Pietro Biroli:** writing - review& editing.

**John Gallis:** resources, data curation, writing - review & editing.

**Joanna Maselko**: conceptualization, writing - review & editing, supervision, funding acquisition.

## Supporting information

S1 Appendix. Flow Chart

S1 Fig. Self-perceptions of ‘being brave’ by response ‘describe the trait among men/women’

S2 Fig. Self-perceptions of ‘being a leader’ by response ‘describe the trait among men/women’

S3 Fig. Self-perceptions of ‘being competitive’ by response ‘describe the trait among men/women’

S1 Table. Characteristics of covariates

S2 Table. Gender attitudes association with standardized outcomes (N=838)

S3 Table. Gender attitudes association with standardized outcomes, complete case (N=810)

S4 Table. Self-perception association with standardized outcomes (N=838)

S5 Table. Baseline variables applied in IPCW modeling among study sample and excluded participants

